# Implementation Toolkit for Small and Sick Newborn Care: bridging the know–do gap through co-design of a global open-access knowledge management web platform and linked community

**DOI:** 10.64898/2026.03.02.26347429

**Authors:** Lauren E Allison, Mbozu Sipalo, Tobias Whatley, Zoe Griffiths, David Gathara, Sarah Murless-Collins, Chinyere Ezeaka, Olufunke Bolaji, Msandeni Chiume, Nahya Salim, Karen Walker, Alex Stevenson, Ralph Hale, Ousmane Ndiaye, Hema Magge, Massimo Salvadori, Federica Cassera, Neena Khadka, Tedbabe Degefie Hailegebriel, Rebecca Richards-Kortum, Maria Oden, Ornella Lincetto, Sara Liaghati-Mobarhan, Harriet Ruysen, Olive Cocoman, Anita Gibson, Gagan Gupta, Joy E Lawn, Implementation Toolkit for Small & Sick Newborn Care Co-Design Group

**Author notes:** Correspondence: Lauren E. Allison. Gagan Gupta and Joy E. Lawn are joint senior authors.

## Abstract

**Background:** Most births worldwide (>80%) occur in health care facilities, yet 2.3 million newborns die annually. If the *know*–*do* gap between evidence and implementation was closed, an estimated 752,000 newborn deaths could be prevented per year. To bridge this gap, we describe the co-design of the Implementation Toolkit for Small and Sick Newborn Care (Newborn Toolkit), a web platform and linked community of global implementers, facilitated by NEST360 and UNICEF. The Newborn Toolkit enables access to practical, curated resources, including tools for peer learning and adaptation to country contexts.

**Methods:** A systematic three step process was followed. Step 1) Structure: We used an organising framework of WHO and UNICEF ten core components for health systems strengthening. We then reviewed relevant knowledge management platforms to identify elements facilitating user engagement. Step 2) Content: >300 implementers collated publications and tools for ten core components. Step 3) Refining and building community: User data analytics and surveys plus direct feedback from the global communities of practice provided data to improve website and webinar content.

**Results:** Step 1) Structure: In 2020, the Newborn Toolkit website structure was co-designed based on the ten core components. Step 2) Content: Working groups, organised by core components, collated over 1,100 resources in 15 languages. Step 3) Refining and building community: Cross-country learning was facilitated through 45 webinars with multi-disciplinary speakers from all continents including caregivers, clinicians, non-governmental organisation representatives, engineers, and data scientists. French language translation and engagement was added between 2023-2025. Unique user counts increased with 28,146 in 2023 to 62,561 in 2025 from 198 countries and territories. The most viewed content includes WHO guidelines, neonatal floor plans, the ABC device costing tool, and data tools.

**Conclusions:** Given the urgency for accelerated progress for newborn survival by 2030, rapid implementation of proven solutions is needed. It is crucial that implementers can access evidence and tools to adapt for their specific context, rather than “reinventing the wheel”. Systems change is complex, requiring novel approaches to make it doable, such as standard, simplified action pathways available on the Newborn Toolkit. Gaps to address include evidence availability in multiple languages.

**KEY FINDINGS:**

1. **WHAT WAS KNOWN?**

- 2.3 million neonatal deaths are estimated annually and 65 countries are at-risk of missing Sustainable Development Goal 3.2 of reducing the number of neonatal deaths to less than 12 per 1,000 live births by 2030
- There exists a *know–do* gap in global newborn health referring to the inability to translate evidence, what is known to work, into practice to improve health outcomes for WHO-UNICEF level-2 Small and Sick Newborn Care
- Frameworks based on country experiences have been created to guide health systems strengthening such as the WHO/UNICEF core components; however, operationalisation is needed
- Evidence, guidelines, and resources for Small and Sick Newborn Care were available; however, there existed a gap in knowledge management platforms dedicated to enabling evidence-based implementation of health systems strengthening interventions in low-resource settings
2. **WHAT WAS DONE THAT IS NEW?**

- We co-designed a knowledge management platform for resources including tools, readings, and case studies to bridge the *know–do* gap for Small and Sick Newborn Care in low-resource settings
- Content and resources on the site were organised according to the ten WHO/UNICEF core components for health systems strengthening plus infection prevention and control
- This platform was operationalised and scaled by linking to, and engaging with, a global community of practice
- We applied a continuous learning and feedback integration approach to develop and refine content, enable accessibility including by language, and tailor engagement initiatives to implementer needs informed by website user and webinar data analytics
3. **WHAT WAS FOUND?**

- The platform hosts over 1,100 tools and readings with resources available in 15 languages
- Between 2021–2025, this platform had 157,452 unique users, from 198 countries and territories
- 45 webinars were hosted between 2022–2025 showcasing implementation case studies and facilitating cross-country learning
- Knowledge management and targeted engagement can enable uptake of information by organising content in practical, digestible, and feasible formats
4. **WHAT NEXT?**

- The *know–do* gap in Small and Sick Newborn Care needs to be closed. Providing up-to-date, relevant evidence that adapts to emerging knowledge and local learning, alongside leadership and financial inputs, could facilitate quality care delivery through data-driven decision making
- Bridging language barriers, through content translation, is a priority and crucial to enable wider access to up-to-date evidence

## BACKGROUND

2.3 million newborn deaths occur annually with sub-Saharan Africa and South Asia accounting for the highest burden. Prematurity is the leading cause of death globally among neonates followed by birth asphyxia/trauma, congenital abnormalities, lower respiratory infections, and sepsis [1]. With appropriate intervention 75% of these deaths can be prevented [2].

The majority of births globally take place in health care facilities; 80% of births in sub-Saharan Africa take place in facilities [3]. Yet many facilities lack adequate capacity to deliver Essential Newborn Care (ENC) required including immediate care at birth, thermal care, resuscitation, breastfeeding, infection prevention, response and recognition to danger signs, and referral [4] as well as critical recommended interventions for level-2 Small and Sick Newborn Care (SSNC), such as assisted feeding, oxygen, and detection and management of neonatal conditions [5]. To reduce preventable deaths through delivery of ENC and SSNC within high burden settings a health systems strengthening approach is required.

The World Health Organization (WHO) and United Nations Children’s Fund (UNICEF), based on learnings from countries that have systemically scaled up SSNC units in the last decade, have developed a framework to guide other countries in the scale up of SSNC with ten core components for health systems strengthening: leadership and governance, financing, human resources, infrastructure, equipment and commodities, data systems, functional referral systems, linkages to maternal care, family and community, and post discharge follow-up [6]. However, criticisms of this framework include a lack of representation of the dynamism and interaction between core components needed for practical implementation [7]. To make health systems strengthening actionable, operationalising this framework is needed, with consideration to the dynamism of health systems change to guide implementation of evidence and quality standards into practice.

A robust evidence base exists to inform essential SSNC interventions; however, uptake of evidence in practice for health systems change is often complex and requires localisation [8]. In global health, the *know*–*do* gap, also called the *theory-practice* gap, refers to the inability to translate evidence, what is known to work, into real world practice [9, 10]. This lack of evidenced-based implementation affects quality of care and ultimately, outcomes. Poor quality care is a bigger burden to mortality reduction than lack of access, with 60% of deaths from conditions amenable to health care due to poor-quality care [8]. Information that is accessible, digestible, and tailored to context is needed for evidence-based decision-making and implementation leading to quality care delivery. Sharing learning between countries and implementers that have grappled with and succeeded to implement sustainably and at scale is vital. Despite the excellent lessons to share, this is not always documented due to local limitations to do so, and if documentation is resourced, there is often a delay, losing time in sharing valuable lessons for implementation in a similar setting or context.

Knowledge management is a means to provide the right information, to the right person, at the right time [11]. Knowledge management encapsulates knowledge generation, storage, processing, transfer, and utilisation [12]. The number of knowledge management platforms for health have increased in recent years with a range of focuses including evidence networks, surveillance tools, observatories, and data platforms [13, 14]. However, there were no knowledge management platforms dedicated to enabling evidence-based implementation for SSNC in low resource settings. We aimed to address this gap through the creation of an online open-access resource platform and linked community — The Implementation Toolkit for Small and Sick Newborn Care, also known as the Newborn Toolkit.

### Aim and objectives

This paper is part of a supplement reporting findings and learnings from Newborn Essential Solutions and Technologies 360 (NEST360), an alliance of partners, including five African governments (Kenya, Malawi, Nigeria, and Tanzania, Ethiopia), working to reduce neonatal inpatient deaths by improving level-2 newborn care in hospitals through device installation, training, and quality improvement. In this paper we aim to describe the cyclical process of co-designing and refining an online open access platform aiming to bridge the *know*–*do* gap in SSNC.

Our objectives were:

**Objective 1**: Review the landscape of existing knowledge management platforms focused on health systems change. Identify and describe key elements and structure that facilitate knowledge organisation and user-friendly engagement with evidence.

**Objective 2:** Co-design platform development and curation of existing evidence for SSNC, organised according to WHO and UNICEF’s core components for health systems strengthening, plus infection prevention and control.

**Objective 3:** Refine content and operationalise engagement with a global community of practice to enable evidence uptake and implementation.

## METHODS

The Implementation Toolkit for SSNC, an open access online resource platform, was developed in collaboration with UNICEF as part of the NEST360 multi-country alliance for neonatal mortality reduction. We adopted a systematic approach comprising three steps: 1) Reviewing existing open access platforms and stakeholder consultations, 2) co-designed platform development, content organisation, refinement, and translation, and 3) deploying engagement initiatives across social networks targeting a global community of SSNC implementers.

### Methods by objectives

#### Objective 1

Platform structure and development was informed by 1) searching for and reviewing existing open access knowledge management platforms relevant to global health and neonatal health topics and 2) consulting a multidisciplinary group of newborn care implementers in low- and middle- income countries. We partnered with a website development company to design and refine the layout of the web platform based on *a priori* criteria for platform structure and content organisation.

**Focus on implementation:** Content on the site needed to facilitate implementation of evidence into practice for level-2 SSNC in low-resource settings. A narrative topic overview was included for each core component on the website. Relevant tools such as guidelines, checklists, and research articles were linked and easily accessible.

**Multi-level systems approach:** A multi-level systems approach was taken to account for inputs to health care delivery beyond the facility level such as national leadership, governance, and financial mechanisms impacting on health delivery. The platform needed to host evidence across all elements of system changes in one accessible location. Organisation was guided by WHO/UNICEF ten core components for health systems strengthening with emphasis on inter-linkages between different core components.

**Neutral & consistent branding:** The platform was branded with a colour palette unique to the Newborn Toolkit and a neutral website domain (www.newborntoolkit.org) with the intention of signalling a collaborative ownership with the global community of implementers and democratising access to evidence.

**Open-access & accessibility:** All learning including content, tools, publications, and webinars needed to be open-access to reduce knowledge sharing barriers and facilitate greater uptake. As the target audience for this platform includes those based in low resource settings, this platform needed to be accessible on mobile and desktop devices and use low bandwidth.

**Ownership and representation:** The platform needed to appeal to a wide range of implementers including policy makers, physicians, nurses, midwives, researchers, funders, caregivers, biomedical engineers, programme managers among others. Mechanisms for community members to give feedback on platform design was a key requirement.

#### Objective 2

Development of the platform and review of potential content occurred in a staged process: 1) content curation and organisation by core component working groups, 2) editorial team revision, 3) Search Engine Optimisation (SEO) and validation, and 4) French language translation.

To achieve this, eleven working groups were established (one for each core component for health systems change plus infection prevention and control), (**Figure 1**) with working group members identified for their related practical experience of implementation and research in this field.

**Figure 1.**
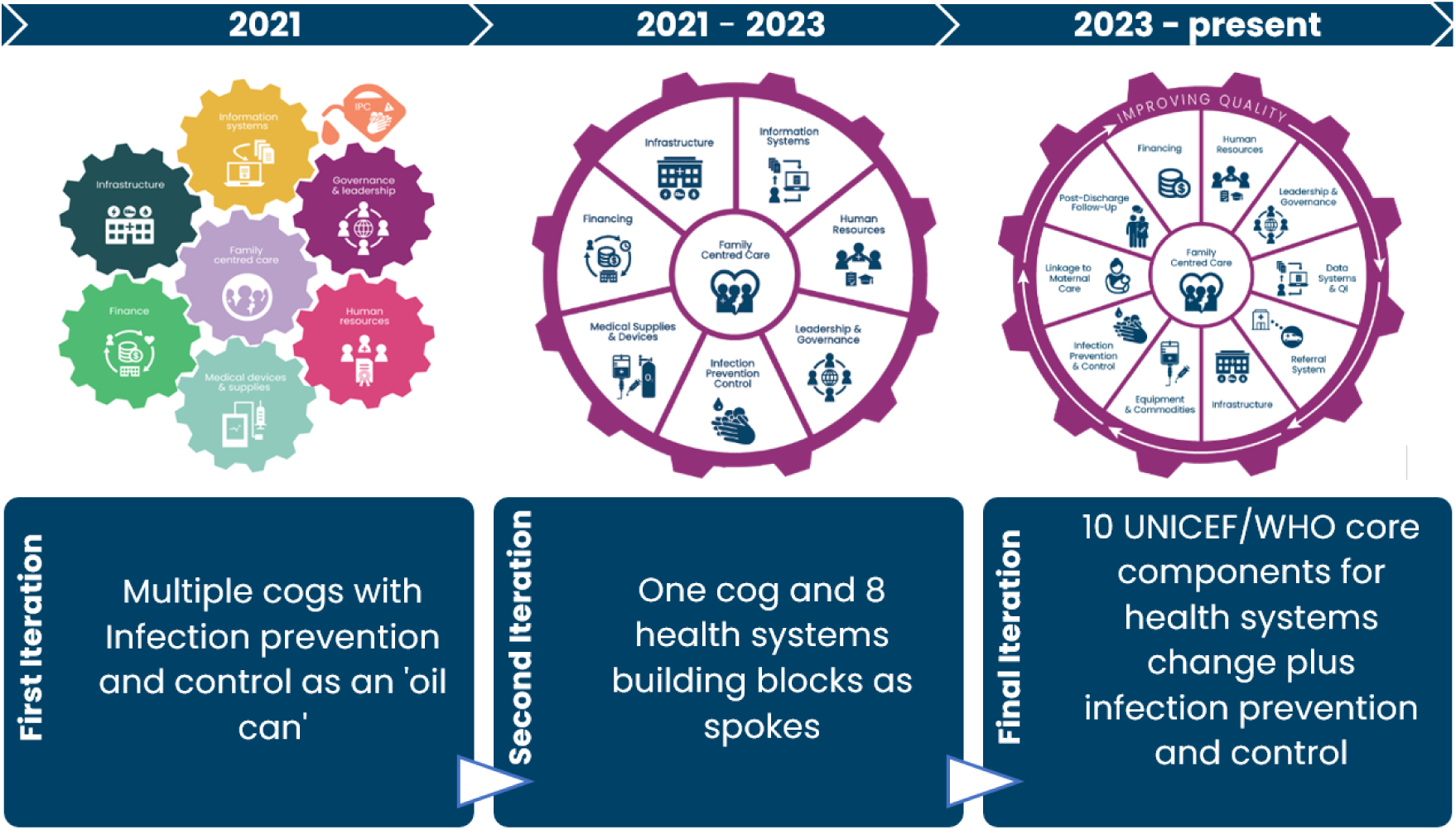
Stages of design of the Newborn Toolkit’s cog incorporating the components for health systems change.

Each of the 11 Newborn Toolkit sections facilitate navigation to reports, checklists, floor plans, strategies, investment cases relevant to that section theme. Working group members identified, screened, triaged, and organised content within their Toolkit section alongside developing a narrative overview of the topic. Content was assessed for inclusion with consideration to quality (peer reviewed, reputable sources), practical guidance, focused on high burden settings for neonatal morbidity and mortality, and in alignment with WHO and United Nations recommendations.

An editorial team comprised of representatives from NEST360 (JEL, ZG, and DG) and UNICEF (GG), ensured messaging and flow were cohesive across content curated by different working groups. A website content coordinator (TW) reviewed and refined all content to ensure it complied with Search Engine Optimisation (SEO) and best web accessibility practices. Content on the site was translated to French using the artificial intelligence-enabled Word Press Multi-lingual (WPML) Plugin. Post-translation review was conducted by a French-language speaker and health care professional to ensure accuracy and meaning were preserved.

#### Objective 3

Engagement with key constituents is necessary for knowledge translation and was central to the Implementation Toolkit for SSNC strategy. Engagement efforts were tailored to reach high-burden settings for neonatal mortality including sub-Saharan Africa and South Asia.

### Engagement Initiatives

Between November 2021 and December 2025, we implemented the following outreach engagement strategies:

A monthly webinar series was created featuring multi-country and multi-lingual learning. Each webinar theme was aligned to one of the ten WHO/UNICEF core components for health systems change. Live translations of webinars were provided from English to French. Webinars followed a systematic format including a topic overview and three examples of local, national, or regional learning across different global regions. Webinars focused on sharing real world examples of implementation underscoring the interaction between health system core components. A question-and-answer period was moderated to enable two-way learning opportunities.

Social media profiles and campaigns were developed for the Toolkit. LinkedIn, Facebook, X (formerly Twitter) and Bluesky were selected as main engagement platforms with a higher presence of relevant professional and caregiver representation.

A monthly newsletter was developed using Mailchimp to share key updates on newborn health and Toolkit-related initiatives. These include announcements of upcoming and recent webinars, new guidance or research, relevant news items, and blogs.

An expert advisory team was created with members representing clinical care, policy makers, funders, researchers, professional associations, and caregivers. Between 2022–2025, this group met quarterly to strengthen inter-disciplinary and cross sectoral collaboration, review engagement activities, and inform the strategic direction of the Toolkit and linked community.

An annual crowdsourcing essay contest was developed by the expert advisory team in 2023 to commemorate World Prematurity Day and promote global discussion on key SSNC themes. Essay topics have focused on making the case for investing in SSNC, advancing family-centred care, and cross-professional neonatal collaboration to improve quality of care.

### Data Analytics

Monthly data analytics reports were developed with indicators to track user engagement and site growth. Website analytics included Google search engine impressions, average search engine position, 28-day active users, new users, route of accessing the site, type of device used to access the site, geographical distribution of users, page views, and length of engagement. Monthly webinar attendance and social media growth and engagement was also monitored.

## RESULTS

### Results by objectives

#### Objective 1

A multi-level knowledge management structure was taken to organise content and resources. The platform was organised using the WHO/UNICEF ten core components for health systems strengthening (plus infection prevention and control) providing a specific landing page for each of the of the core components.

The main body of each core component contains narrative style topic overview and sub-sections with links to related publications and tools enabling easy access to resources. Tools and readings can be filtered according to language, publication year, and topic. Topics are organised into UN Guidelines, Single Country Evidence, and Multi-Country Evidence.

### Design and Toolkit Cog

Several iterations of the platform visual framework were developed (**Figure 1**). A cog was selected as the main shape to reflect the dynamism and interrelatedness between the core components. A second iteration was developed as a circular cog with a seamless flow between cog spokes reflecting eight core components for health systems change plus infection prevention and control. The final version of the cog was updated in 2023 to reflect the WHO/UNICEF ten core components of health systems strengthening plus infection prevention and control acting as ‘spokes’ in the cog. Family-centred care was placed in the middle to emphasise the need for partnership and co-design with caregivers throughout the process of systems change. A unique colour palette was selected for the Newborn Toolkit branding to facilitate visibility and branding independent of NEST360 and UNICEF.

#### Objective 2

As of December 2025, the site hosts a total of 585 tools and 524 readings. Among these resources, 132 tools and 125 readings are available in French. The Toolkit hosts many multilingual tools that have been translated and are available in up to 15 languages.

The health systems core component sections contain a narrative overview of the topic and core themes outlining essential steps to improve health delivery in the context of SSNC. Each core component follows the same general layout with an overview section and then 3–8 sub-sections for key themes (**Figure 2**).

**Figure 2.**
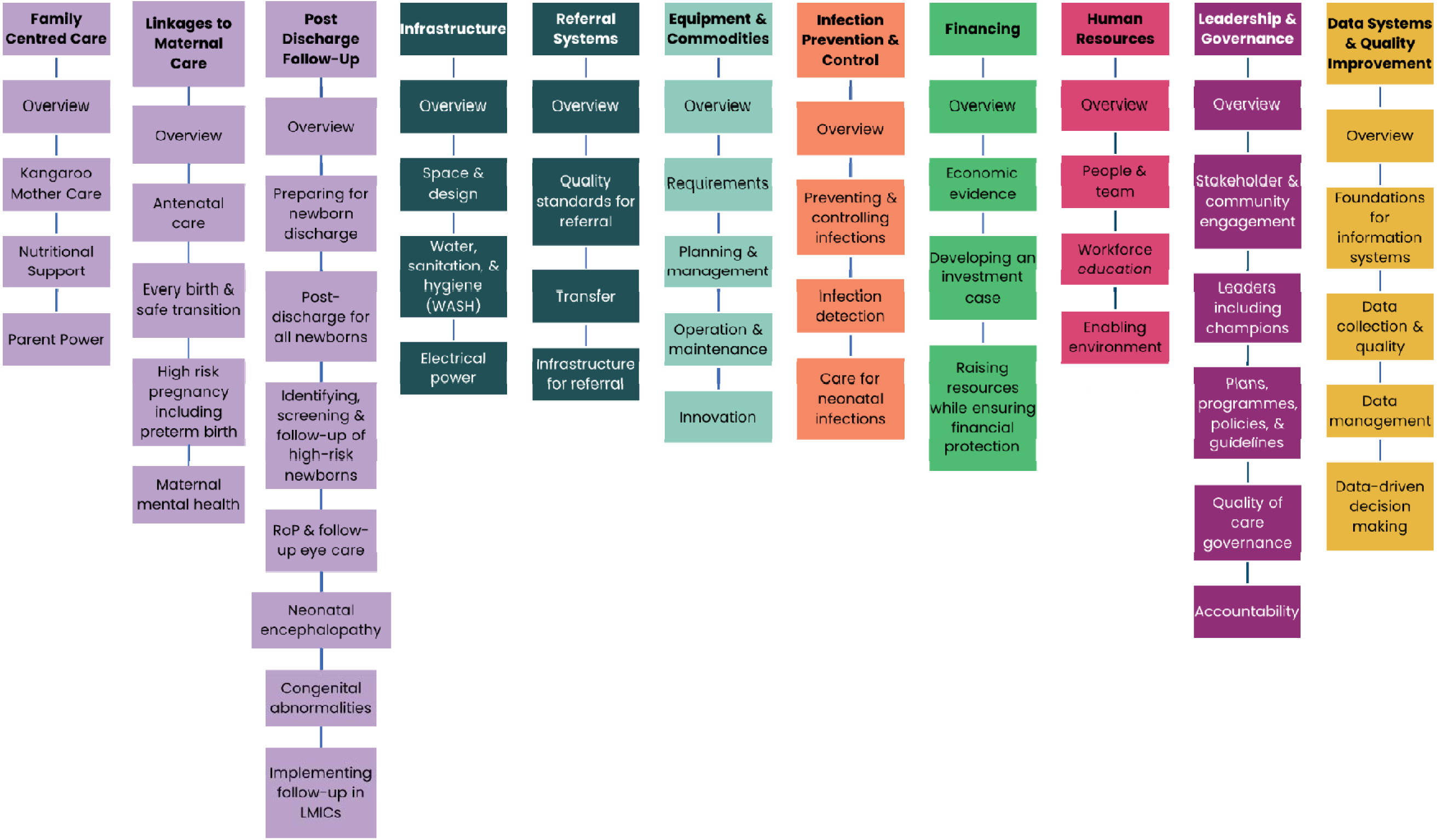
Website technical content organised according to health systems core components plus infection prevention and control.

Between 2022–2025, among all health systems core components the most accessed include the following: Data Systems and Quality Improvement, Equipment and Commodities, Infrastructure, Human Resources, and Infection Prevention and Control (**Table 1**).

**Table 1.** Most accessed resources and tools by WHO and UNICEF core components for health systems strengthening, 2022–2025.

| 2022 | 2023 | 2024 | 2025 |
| --- | --- | --- | --- |
| Human Resources | Infrastructure | Equipment & Commodities | Data Systems & Quality Improvement |
| Quality Improvement | Data Systems & Quality Improvement | Infection Prevention & Control | Infrastructure |
| Leadership & Governance | Equipment & Commodities | Infrastructure | Equipment & Commodities |

Most accessed resources between 2022–2025 included WHO guidelines, neonatal unit floor plans, and NEST360 resources (**Table 2**). This includes WHO standards for improving quality of care for small and sick newborns in health facilities, the Tanzania Regional hospital neonatal floor plan, and the ABC Neonatal Devices Planning and Costing Tool developed by NEST360. Country specific tools were also commonly accessed including tools with a contextual framing in India. This includes The India Newborn Action Plan, India complete sample case record forms, and India: Facility Based Newborn Care (FBNC) Facilitator Guide among others. In 2024, NEST360 tools and resources grew in popularity including NEST360 BMC Supplement *— Small and sick newborn care: learning for implementation across Africa and beyond*, NEST360 Health Facility Assessment (HFA) Paper Tools, and NEST360 Facility Quality Improvement Dashboard.

**Table 2.**
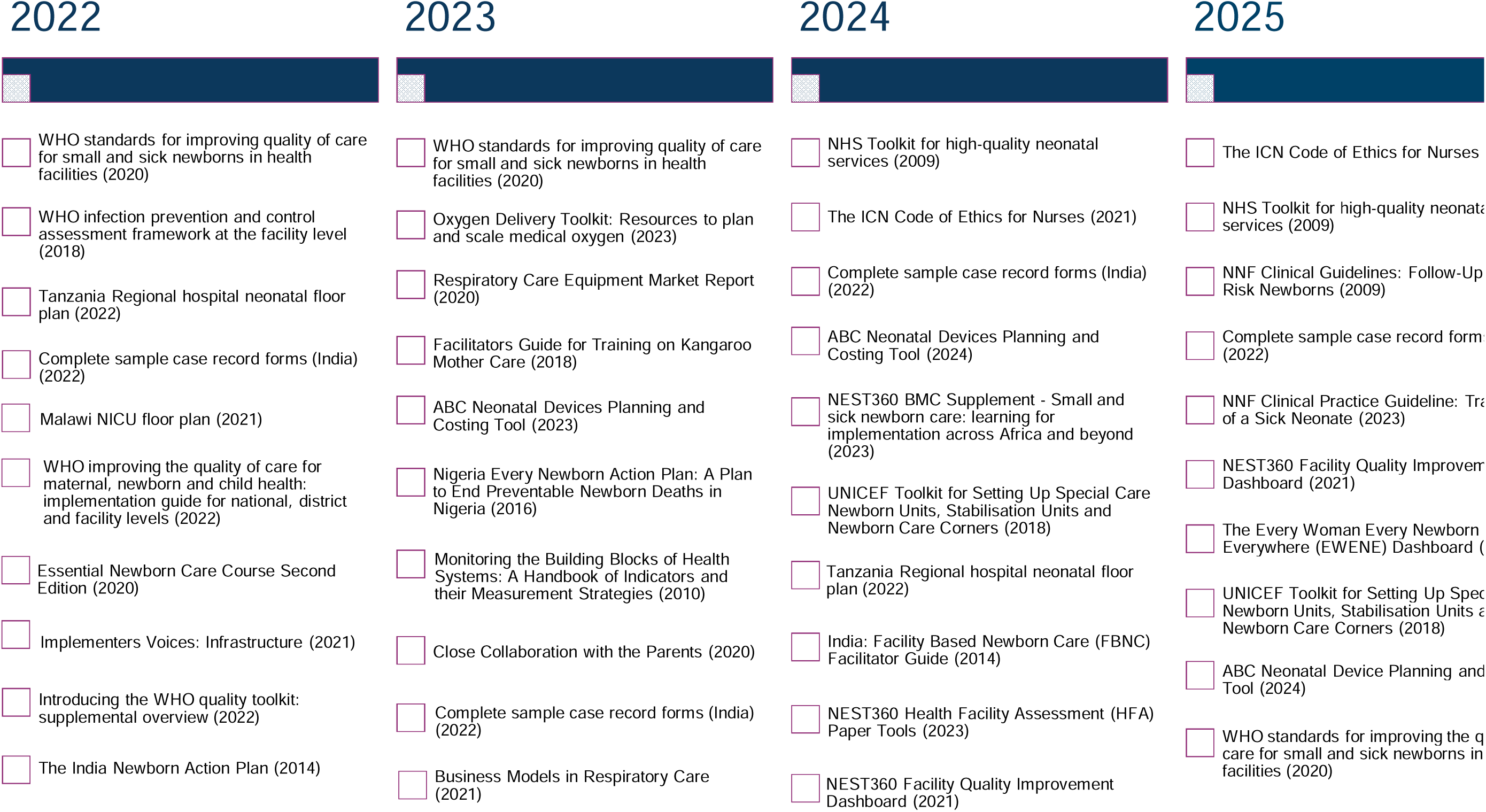
Most accessed resources on the Newborn Toolkit by year, 2022–2025.

In 2024, to simplify the complexity of systems change and support identification of relevant resources, the Newborn Toolkit developed implementation action pathways. These action pathways include a step-wise process to facilitate implementation of evidence into practice with a specific topic focus. Key tools to support implementation are linked along relevant points in the pathway (**Figure 3**). Pathway themes include Pain Prevention and Management, Quality Kangaroo Mother Care, Neonatal Unit Outbreak Response, and Jaundice Management.

**Figure 3.**
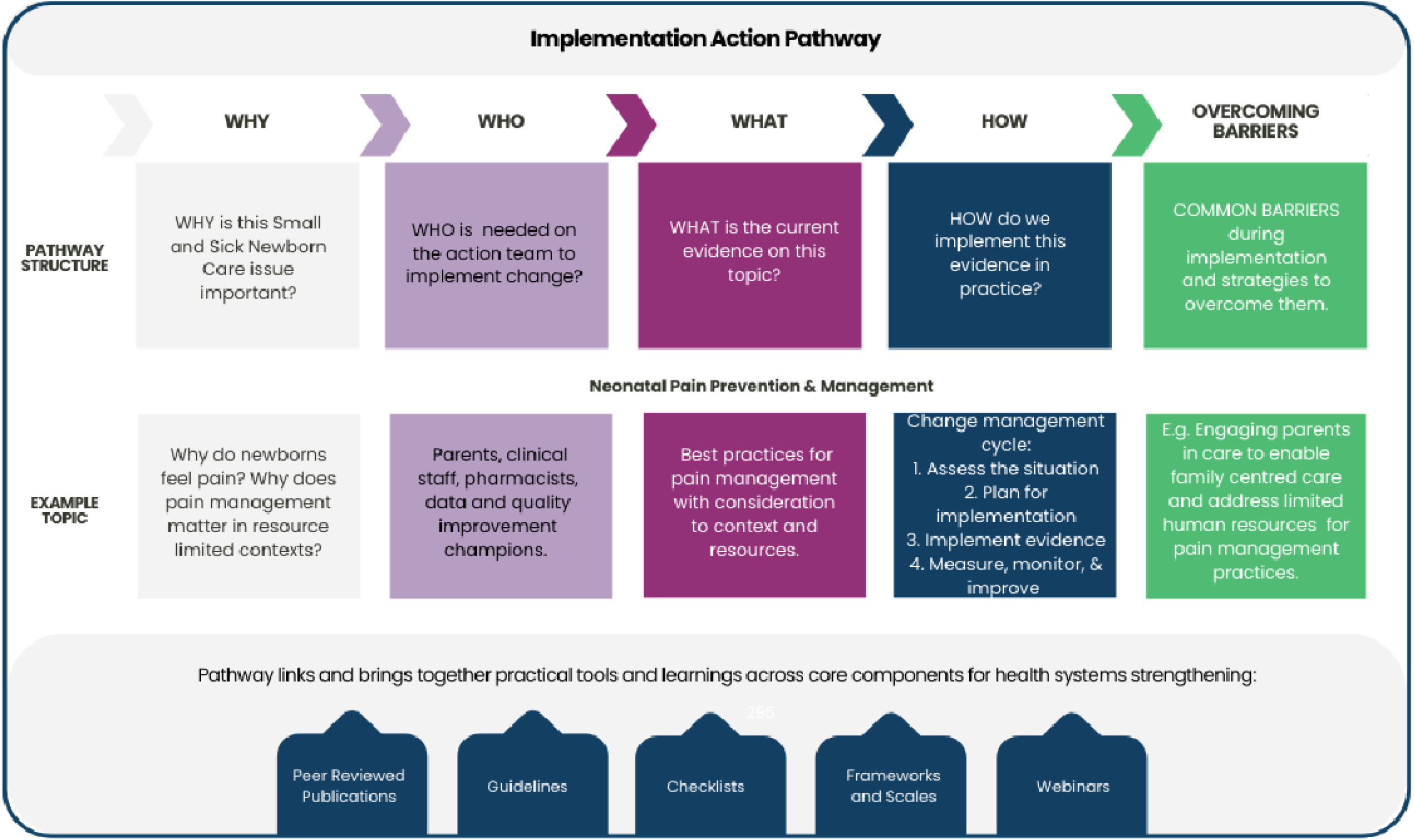
Implementation action pathway structure and neonatal pain prevention and management example.

#### Objective 3

The platform called the Implementation Toolkit for SSNC, or Newborn Toolkit, was launched publicly on November 17, 2021 linked to World Prematurity Day.

Between November 2021 and December 2025, 157,452 unique users accessed the site (**Figure 4**). Unique user growth increased year on year with 10,273 in 2022, 28,146 in 2023, 54,059 in 2024, and 62,561 in 2025 (**Figure 4**). October and November in 2023, 2024, and 2025 demonstrated major increases in users with a lower number of users accessing the site in January and February.

**Figure 4.**
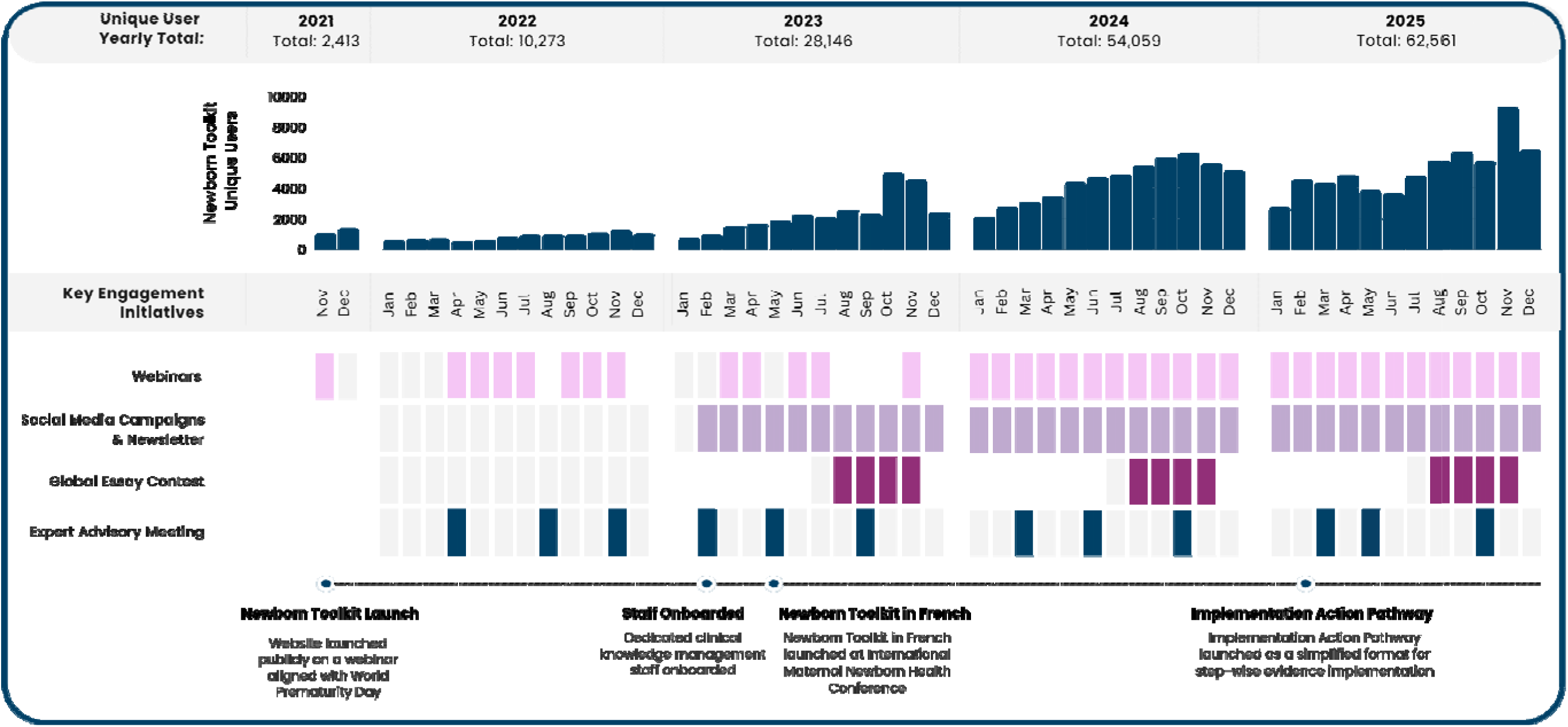
Newborn Toolkit monthly website unique user count and frequency of key engagement initiatives, November 2021 – December 2025.

As of 2025, users accessed the site from over 198 countries and territories. Between 2022–2025, among all countries (**Figure 5**), the following countries were identified as where the Toolkit was most accessed: United States, India, China, United Kingdom, Kenya, Nigeria, Tanzania, Australia, Netherlands and Uganda. The top World Bank classified LMIC countries (**Figure 6**) where the Toolkit was accessed from include India, China, Kenya, Nigeria, Tanzania, Uganda, South Africa, Ethiopia, Malawi, and Ghana. The top French-speaking countries where the Toolkit was accessed include the following: France, Democratic Republic of the Congo, Rwanda, Cameroon, Senegal, Burkina Faso, Switzerland, Ivory Coast, Benin, and Burundi. Among high-income countries the Toolkit was most accessed from United States, United Kingdom, Australia, Netherlands, Singapore, Ireland, France, Canada, Italy, and Germany.

**Figure 5.**
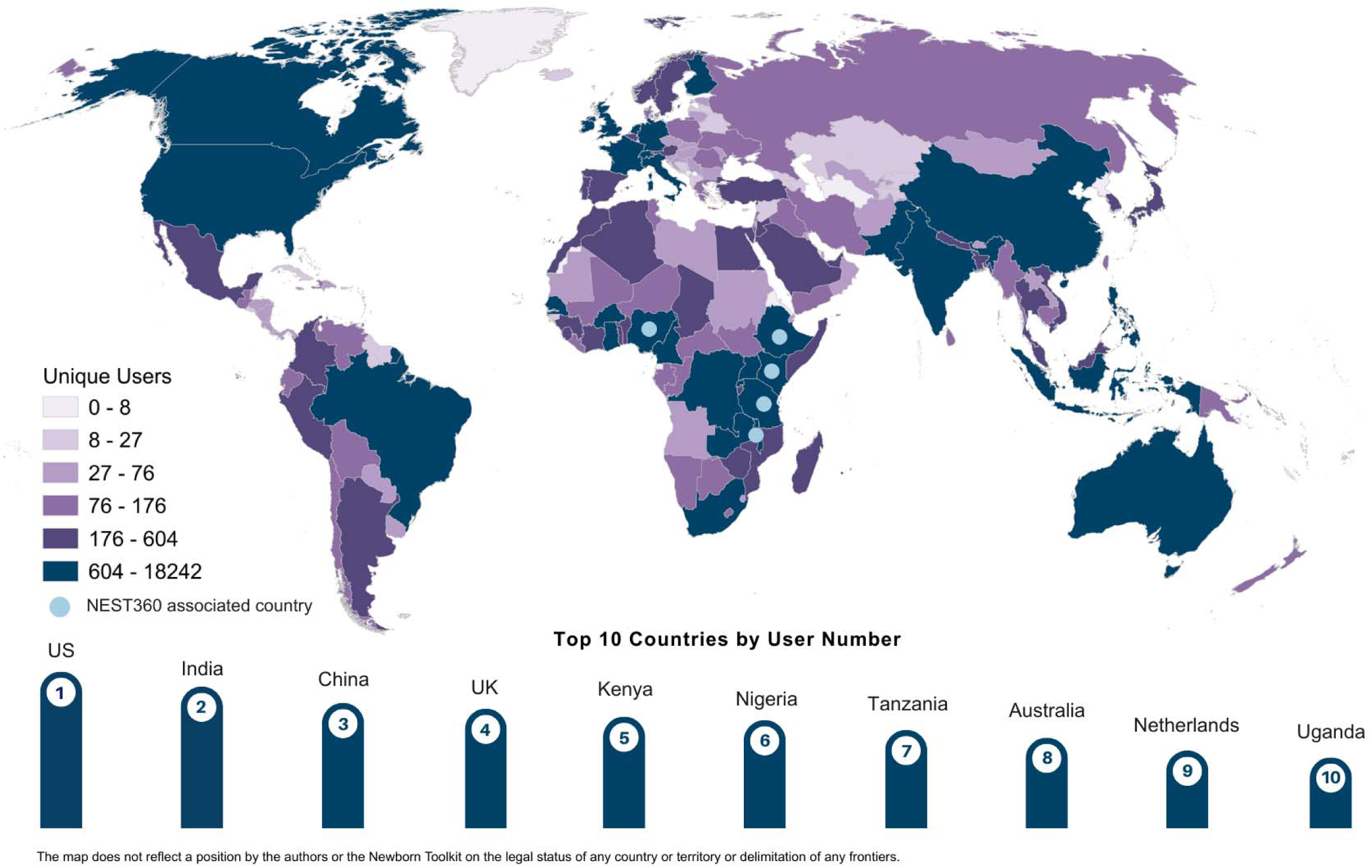
Global distribution of Newborn Toolkit active users, 2022–2025.

**Figure 6.**
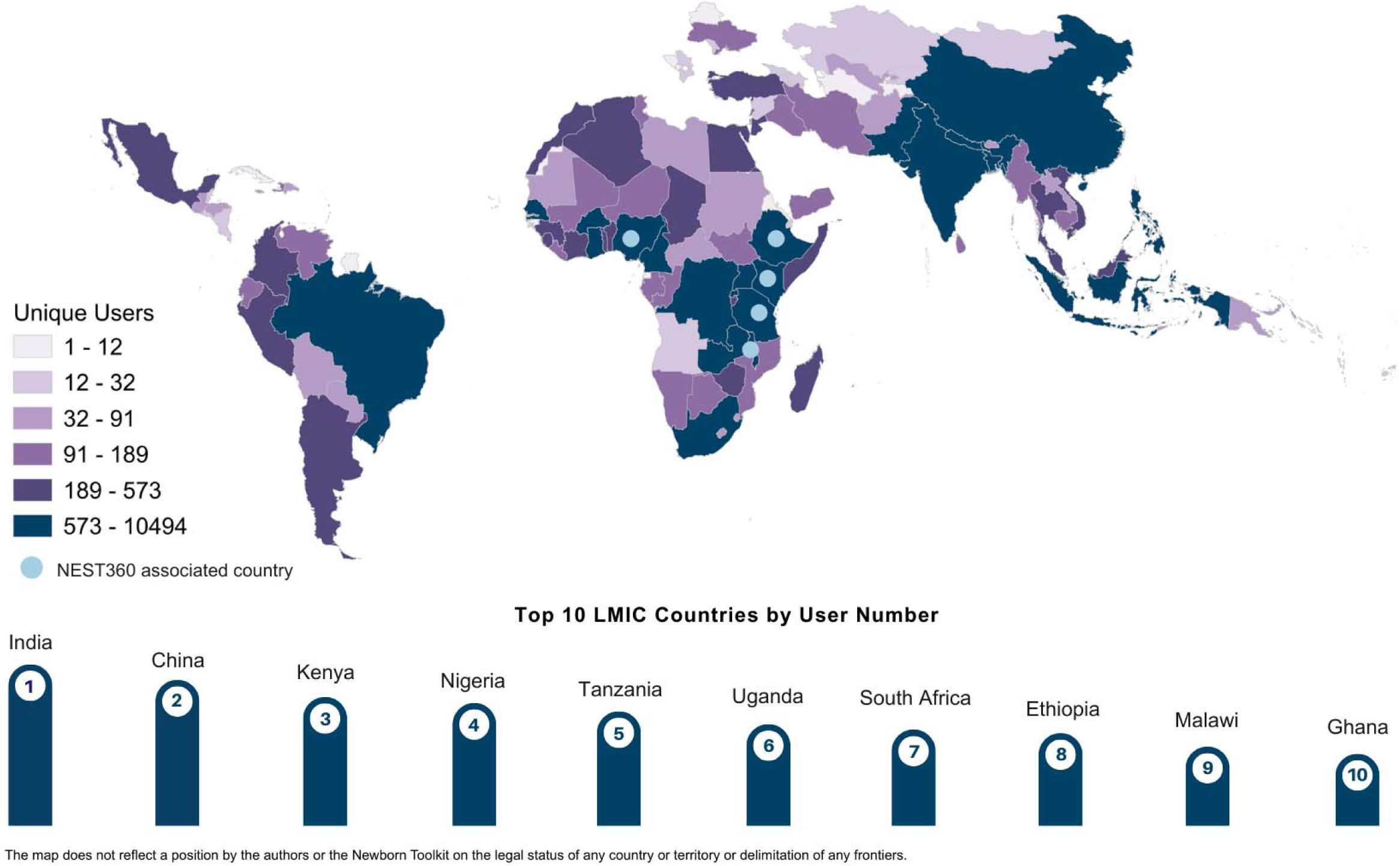
Global distribution of Newborn Toolkit active users among World Bank classified Low- and Middle-Income Countries, 2022–2025.

Since November 17, 2021, 45 webinars have been hosted by the Newborn Toolkit, aligned with the WHO-UNICEF health system core components. Forty of these webinars have been delivered in English with live French translation. Three webinars were delivered in French with one additional webinar delivered in Spanish. In 2024 alone, the webinars reached nearly 3,500 viewers, through both live sessions and catch-up recordings. Among all webinars, the highest in attendance included 1) What’s next for Kangaroo Mother Care before clinical stabilisation, with over 280 live attendees; 2) Kangaroo Mother Care with quality: duration, nutrition, & data that matter with over 150 live attendees and 3) Babies and bugs: Innovations to address neonatal sepsis and antimicrobial resistance, with over 120 live attendees. Speakers represented caregiving, clinical, academic, non-governmental, governmental, and private sector backgrounds. This includes WHO, UNICEF, Pan American Health Organization, International Paediatric Association, Council of International Neonatal Nurses, Clinton Health Access Initiative, Global Foundation for the Care of Newborn Infants among others.

## DISCUSSION

This paper describes the development of a web platform and linked community of practice for evidence implementation of SSNC. The Newborn Toolkit content and community engagement approach is a continuous process informed by regular user feedback and in alignment with data on global newborn health. Continuous inputs have enabled the Newborn Toolkit to adapt and pivot to address current needs of implementers of SSNC in high burden settings. This process has shaped the Newborn Toolkit into an interprofessional educational platform that engages those implementing SSNC globally, with 157,452 unique users from 198 countries and territories between 2021–2025.

### Global distribution of users and practical challenges to uptake

Among low- and middle-income settings, the Toolkit has an active user base in high burden settings including sub-Saharan Africa, South Asia, and South-East Asia [1]. Top countries where users accessed the Newborn Toolkit include the five countries (Kenya, Malawi, Tanzania, Ethiopia, Nigeria) where NEST360, comprising clinicians, biomedical engineers, facility management, and policy makers, is working to improve essential SSNC delivery in over 130 health facilities. The Newborn Toolkit contains many implementation research findings arising from the work undertaken by NEST360 [15, 16]. In the context of managing health conditions, local knowledge production promotes greater sense of ownership, trust, and usable findings that are relevant to context [17]. The linkages between the Toolkit and evidence produced in NEST360-linked health facilities may be an enabler of high engagement among country users. Fostering and leveraging similar partnerships whereby the Newborn Toolkit can host and actively partner to elevate local learning could be an enabler for engaging implementers in other high burden contexts.

Although the target audience of the Newborn Toolkit is for SSNC implementers in low-resource settings, the platform still generates significant interest among high-income countries. Country population size, increased neonatal healthcare professionals, non-governmental organisations, and researchers, may contribute to number of unique users accessing the site by country. Notably, users based in India and the United States were top website users. Both countries have comparatively large populations and health workforces [18, 19]. High-income countries such as the United Kingdom and Australia have bigger neonatal care workforces with lower patient ratios [20, 21].

The Toolkit platform is designed to use low bandwidth and permits users to save offline PDF files; however, certain low-resource settings experience added challenges with power shortages, limited internet infrastructure, and lack of information technology support [22] further impeding engagement with the Newborn Toolkit.

### Engagement strategies

With lower platform user engagement initially, significant growth was noted between 2022 to 2025. Growth was enabled by addition of two technical staff including an African neonatal medical doctor and a bilingual English/French neonatal nurse. Technical staff led the community of practice engagement initiatives from 2023 onwards. Fluctuations in website users were generally aligned with Newborn Toolkit engagement activities. Notably, October and November tended to have higher engagement rates; the Newborn Toolkit hosted an annual crowdsourcing essay contest leading up to World Prematurity Day engaging grass-roots implementers of SSNC. Monthly webinars and promotion of the Newborn Toolkit at related conferences also tended to increase unique user engagement.

A strength of the Newborn Toolkit webinars is the diversity in topics. Varied webinar topics and speakers also attracted a diverse viewership with different webinar topics tailored to and more appealing to different disciplines. For instance, a webinar sharing interdisciplinary perspectives on devices offered practical guidance for biomedical engineers while a webinar on nursing qualifications was targeted towards a nursing audience. Targeted engagement also has important implications for evidence uptake. Notably, nurses reported lack of local contextual evidence as major barriers to evidence implementation [23]. Similarly, a review found that policy makers reported barriers to evidence uptake as a lack of access to good quality relevant research [24]. Other factors limiting evidence uptake included poor dissemination of research findings and limited research skills of evidence implementers [23, 24]. The Newborn Toolkit webinars offer a medium to integrate role specific considerations when presenting evidence and may be better able to frame evidence for uptake among different audiences with varying research skills.

### Engaging a multilingual community

In response to the large representation of French-speaking countries among high burden settings [1], French translation of the Newborn Toolkit content and initiatives was identified as a priority with funding secured in 2023. However, language limitations were noted in the availability of tools and readings. English remains the dominant language of science communication [25, 26]. Compared to over 800 tools available in English, there are only 250 tools in French on the Toolkit. Delays or lack of publication of key evidence in non-English languages create barriers for non-English speakers in accessing and implementing up-to-date evidence [27]. Notably, French and Portuguese speakers based in Africa reported preference for publishing in English as there are more higher impact journals with larger readerships [28]. Similarly, to increase language accessibility, the Newborn Toolkit hosted webinars featuring French only speakers; however these webinars were poorly attended by French and English speakers. Webinars featuring English speakers or hybrid English/French speakers were better attended by French-speakers compared to webinars featuring only French-speakers. This preference for English-speaking webinars could be attributed to the webinar topics with topic selection often guided by latest evidence related to SSNC. This evidence is often published first in English and thus presented and discussed by English-speakers on webinars.

The Toolkit narrative overview content and engagement initiatives are currently only offered in English and French. Within Africa alone, there are over 1,000 languages spoken with many regional dialects [28]. Fewer languages and limited tailoring to regional cultural norms may limit the uptake of the Toolkit particularly among those speaking minority languages. Translation to additional languages including Portuguese and Spanish is a priority focus. Advancements in AI-enabled translation permit faster translation and accessibility, however limitations in the accuracy of medical evidence translation, may pose ethical and safety challenges [29, 30], particularly in the context of vulnerable newborns. To bridge this language divide, journals and organisations that produce health guidance could consider translation of evidence to non-English languages, with post-translation review, particularly for findings that have potential for high impact in non-English speaking settings.

### Simplifying complexity

Organisation and delivery of content have important implications for uptake and use of evidence in practice. Enablers of engagement linked to website design include ease of navigation, presence of graphics, website organisation, content utility, purpose, simplicity, and readability [31]. In the context of health guidance, a review identified that challenges in evidence uptake were linked to lack of guideline availability, difficultly in locating guidance, lack of staff training on guidance updates, and multiple different international and national guidelines [32]. Establishing and improving quality of care delivery across multiple components of a health system is a complex process that requires nuance stemming from a large body of evidence [33]. A strength of the Newborn Toolkit is its robust repository of evidence for quality care delivery, located in one convenient place. However, with over 1,100 resources, for users, it can be overwhelming to discern which tools and evidence are best. Implementation Action Pathways provided a means to support uptake of evidence in a simplified step-wise approach. These pathways also identified varied target users and integrated implementation considerations for different health professionals into the approach for applying evidence in practice.

### Strengths and Limitations

A strength of the Newborn Toolkit includes the systematic development process including continuous feedback incorporated to target gaps and guide website improvement. The Toolkit is a dynamic platform that is continuously being updated and is flexible and adaptable to emerging guidance. Development of the website content was guided by a framework, WHO/UNICEF’s health systems ten core components, and was informed by input from over 300 implementers of SSNC honing a breadth and depth of topic experience. The Toolkit hosts a large repository of practical tools in over 15 languages.

A limitation includes the inability to measure the impact of the Toolkit directly on user knowledge change and implementation. It is challenging to quantify the direct effects of how the data is being used at the health facility level. Beyond engagement with the Toolkit, implementers require many inputs at international, national, and local levels to achieve meaningful health systems improvement. Additional evaluation is needed to assess the impact of the Newborn Toolkit along the continuum of evidence implementation. A further limitation is the sustainability of such platform which requires dedicated human and financial resourcing, and fundraising for improvements.

The Newborn Toolkit increased engagement activities significantly during 2023 facilitated by expansion of staff with a focus on knowledge management and clinical care. Lower unique users in 2022 may be linked to less engagement and exposure opportunities rather than user interest. Data from 2021 was excluded from analysis of site analytics for most common tools and resources accessed as the website was launched publicly in November 2021, leading to a smaller less representative sample size during 2021.

### Conclusions

With 2.3 million neonatal deaths estimated annually, and 65 countries off-track to reaching the Sustainable Development Goals of less than 12 newborn deaths per 1,000 live births by 2030, progress on SSNC needs to be accelerated. By providing up-to-date, relevant evidence and local learning, alongside leadership and financial inputs, the Newborn Toolkit facilitates quality care delivery through data-driven decision making. Expanding accessibility and uptake of evidence in high-burden settings is a priority to enable level-2 SSNC. To achieve this, policies should focus on enabling increased language offerings of evidence, locally led content development, and enabling opportunities for cross-country learning with consideration to context and resources.

## LIST OF ABBREVIATIONS

LMIC: Low- and Middle-Income Country
LSHTM: London School of Hygiene & Tropical Medicine
NEST360: Newborn Essential Solutions and Technologies 360
ROP: Retinopathy of Prematurity
SEO: Search Engine Optimisation
SSNC: Small and Sick Newborn Care
UN: United Nations
UNICEF: United Nations Children’s Fund
WHO: World Health Organization
WPML: Word Press Multilingual

## DECLARATIONS

### Ethics and consent to participate

Ethical approval was not sought, as no clinical or service data were obtained from stakeholders during development of the Implementation Toolkit for Small and Sick Newborn Care.

### Consent for publication

Not applicable.

### Availability of data and material

The dataset is available in the public domain on the Implementation Toolkit for Small and Sick Newborn Care (https://newborntoolkit.org/).

### Competing interests

The authors declare that they have no competing interests.

### Funding

This work is funded through the NEST360 Alliance with thanks to the John D. and Catherine T. MacArthur Foundation, Gates Foundation, ELMA Philanthropies, The Children’s Investment Fund Foundation (CIFF), The Lemelson Foundation, Sall Family Foundation, and the Ting Tsung and Wei Fong Chao Foundation under agreements with William Marsh Rice University.This work is also funded by the UK Research and Innovation, Medical Research Council and the Paolo Chiesi Foundation.

### Authors’ contributions

This work was done in partnership with the NEST360 Alliance and the Implementation Toolkit for Small and Sick Newborn Care Co-Design Group. The Implementation Toolkit for Small and Sick Newborn Care was conceptualised by the NEST360 Alliance facilitated by JEL, DG, and GG. LEA and JEL developed the objective framework and methodology for this paper. The framework and methodology underwent additional refinement with inputs from co-authors. Website analytics were provided by TW. The original manuscript was drafted by LEA. Manuscript graphics were created by LEA. Reviewing and editing was undertaken by all authors. All authors reviewed and gave their consent to the final version of the manuscript. The authors’ views are their own, and not necessarily from any of the institutions they represent.

## Acknowledgements

Firstly, and most importantly, we thank the women, families, and newborns together with all the health workers involved in this work. We also thank those involved as part of the core component working groups named under the Implementation Toolkit for Small and Sick Newborn Care Co-Design Group. Many thanks to Roisin Henry for administrative and operational support. During the study period, JEL, LEA, MS, TW, ZG, DG are partially funded through the NEST360 Alliance, with thanks to the John D. and Catherine T. MacArthur Foundation, the Bill & Melinda Gates Foundation, ELMA Philanthropies, The Children’s Investment Fund Foundation UK, The Lemelson Foundation, and the Ting Tsung and Wei Fong Chao Foundation, under agreements to William Marsh Rice University. LEA is also partially funded by the Paolo Chiesi Foundation. LEA and MS are also partially funded by UKRI Medical Research Council. Finally, we are grateful to fellow researchers and guest editors who reviewed this paper, and for the input from managing editors at BMC and within NEST360 including Kristina Shemwell, William Macharia, Caroline Noxon, and Joy E. Lawn.

## Implementation Toolkit for Small and Sick Newborn Care Co-Design Group

Group members are displayed according to the WHO/UNICEF core component website section for which they contributed content development and curation.

**Data Systems and Quality Improvement**: Aeesha NJ Malik, Eric Ohuma, Gagan Gupta, Geoffrey Manda, Ifeanyichukwu Anthony Ogueji, Irabi Kassim, John Wainaina, Josephat Mutakyamilwa, Josephine Shabani, Joy E. Lawn, Karen Walker, Karim Manji, Louise Tina Day, Msandeni Chiume, Nahya Salim Masoud, Olabisi Dosunmu, Rebecca Penzias, & Samuel Ngwala.

**Leadership and Governance**: Alex Stevenson, Carole Kenner, Chinyere Ezeaka, Donat Shamba, Evelyn Zimba, Fatima Gohar, Gaurav Sharma, Karen Walker, Mary Kinney, Meghan Kumar, Richard Kagimu, & Timothy Powell-Jackson.

**Human Resources**: Anniina Lockwood, Carole Kenner, Charles Osuagwu, Chinyere Ezeaka, Edith Gicheha, Ekran Rashid, Ephraim Kumi Senkyire, George Banda, Goldy Mazia, Grace Tahuna Soko, Jennifer Werdenberg-Hall, Josephat Mutakyamilwa, Ifeanyichukwu Anthony Ogueji, Karen Walker, Karim Manji, Meghan Kumar, Merran Thomson, Neena Khadka, Olufunke Bolaji, Richard Kagimu, Ruth Davidge, & Sara Liaghati-Mobarhan.

**Equipment and Commodities:** Charles Osuagwu, Elizabeth Asma, Elizabeth Ngowi, George Banda, Hamish Graham, James H. Cross, Jennifer Werdenberg-Hall, Joseph Bailey, June Madete, Maria Oden, Megan Heenan, Patricia Coffey, Rebecca Richards-Kortum, Robert Tillya, Ryan Johnston, Sara Liaghati-Mobarhan, Sarah Collins, & Taylor Boles.

**Financing:** Alison Morgan, Meghan Kumar, Prarthna Dayal, & Opeyemi Odedere.

**Infrastructure:** Chris Harnish, Christine A. Bohne, Donat Shamba, Georgina Msemo, Hamish Graham, Harish Chellani, Ifeanyichukwu Anthony Ogueji, Karim Manji, Kiersten Israel-Ballard, Kondwani Kawaza, Megan Heenan, Nahya Salim Masoud, Natalie Dottle Mitchell, Padraic Casserly, Prateek Gupta, & Sara Liaghati-Mobarhan.

**Family Centered Care:** Aeesha NJ Malik, Alex Stevenson, Carole Kenner, Clare Gilbert, Edith Gicheha, Ephraim Kumi Senkyire, Grace Tahuna Soko, Harish Chellani, Ifeanyichukwu Anthony Ogueji, Jaya Chandna, Josephat Mutakyamilwa, Karen Walker, Karim Manji, Kiersten Israel-Ballard, Louise Tina Day, Maureen Majamanda, Neena Khadka, Rosie Steege, Ruth Davidge, Silke Mader, & Tedbabe Degefie Hailegebriel.

**Infection Prevention and Control:** Ephraim Kumi Senkyire, Elizabeth Molyneux, Gaurav Sharma, Grace Tahuna Soko, Ifeanyichukwu Anthony Ogueji, James H. Cross, Joy E. Lawn, Kalpana Upadhyay Subedi, Karim Manji, Kondwani Kawaza, Nahya Salim Masoud, Naomi Spotswood, & Sarah Collins.

**Linkages to Maternal Care:** Caroline Kinsella, Laura Fitzgerald, Lauren E. Allison, Neena Khadka, Robyn Churchill, Shanon McNab, & Suzanne Stalls.

**Referral Systems:** Aduragbemi Banke-Thomas, David Gathara, Lauren E. Allison, Loveday Penn Kekana, & Peter Acker.

**Post-Discharge Follow-Up:** Cally Tann, Jaya Chandna, Mbozu Sipalo, & Samantha Sadoo.

## Notes

### Competing Interest Statement

The authors have declared no competing interest.

